# The Design of Lil’Flo, a Socially Assistive Robot for Upper Extremity Motor Assessment and Rehabilitation in the Community Via Telepresence

**DOI:** 10.1101/2020.04.07.20047696

**Authors:** Michael J Sobrepera, Vera G Lee, Michelle J Johnson

## Abstract

**Introduction:** We present Lil’Flo, a socially assistive robotic telerehabilitation system for deployment in the community. As shortages in rehabilitation professionals increase, especially in rural areas, there is a growing need to deliver care in the communities where patients live, work, learn, and play. Traditional telepresence, while useful, fails to deliver the rich interactions and data needed for motor rehabilitation and assessment.

**Methods:** From prior work, we have developed design requirements for a socially assistive robot for upper extremity motor assessment and rehabilitation via telepresence. We designed Lil’Flo, targeted towards pediatric patients with cerebral palsy and brachial plexus injuries. The system combines traditional telepresence and computer vision with a humanoid, who can play games with patients and guide them in a present and engaging way under the supervision of a remote clinician.

**Results:** The humanoid’s arms have sufficient range of motion, and the face is able to communicate several emotions clearly. The system is portable, extensible, and cheaper than our prior iteration. A simple web interface allows operators to focus on interactions while the computer vision system stores data for analysis.

**Conclusions:** Lil’Flo represents a novel approach to delivering rehabilitation care in the community while maintaining the clinician-patient connection.

## Introduction

There is a growing need to provide motor rehabilitation in rural communities and other resource-scarce areas. Many patients in rural areas are currently underserved by physical and occupational therapists ^1,2^. Due to a lack of local therapists, these patients must travel long distances to reach a center of excellence to receive care. For patients, this requires a caretaker, often a family member, to take time off work or take time off school. There is a need to improve care for both pediatric and adult patients who reside in rural areas; we focus on the pediatric patients in this work. Two large pediatric patient population that are in need of life long rehabilitation are cerebral palsy (CP) and brachial plexus birth palsy (BP). CP is the most common motor disorder in young children, occurring in 2–3 per 1000 live births ^3^. It results in motor disorders of the upper and lower limbs that are often accompanied by other impairments in sensation, perception, cognition, communication, and behavior ^4,5^.

Brachial plexus birth palsy, also known as perinatal brachial plexus injury, obstetric brachial plexus injury, and simply brachial plexus (BP) has a different mechanism and pathology than CP, but similarly affects upper extremity function. BP occurs as a result of damage to the brachial plexus nerves during delivery, and occurs in about 1 per 1000 births ^6^. Outcomes from brachial plexus injuries can vary from self-resolving to long-term disability, requiring rehabilitation, potential surgery, and long-term management ^7^.

CP and BP patients need rehabilitation care and medical services beyond the hospital, in community-based settings. In addition to local clinics, pediatric patients may have some form of care in their school or a local primary care clinic. In a specialized school, they may have access to an in-house therapist; elsewhere, they may have access to a traveling therapist who visits them for a short period each week. The remainder of their rehabilitation is often done at home with the aid of family members.

Rehabilitation is often a long-term need for CP and BP patients, and as a result compliance and motivation is often an issue. A lack of compliance may lead to decreased functional outcomes. Although compliance can be monitored when the child is with a therapist, it is more challenging at home. Routine diagnostics and assessments are critical to ensure that patients are receiving the best treatment for their condition and current state ^8^. This is especially important for rehabilitation, where patients with the same pathology can have divergent manifestations and challenges in their daily lives ^9^. Frequent assessment of function can give the care team a good indication of a patient’s progress and could provide a method to motivate patients, and is therefore an important aspect of ongoing rehabilitation.

One way to address clinician shortages in medically underserved areas is through telemedicine: the remote application of medicine using telecommunications, which generally include two-way video and audio ^10^. For example, evidence has shown that range of motion in adolescents can be measured effectively via telemedicine, and subjects may prefer telemedicine visits for certain tasks ^11^. For clarity, we define telemedicine as specifically using remote audio and video (i.e. telepresence) for medicine and allow electronic health care (e-health) to define the larger scope of any medicine practiced using remote electronic communications. Telemedicine enables patients to receive care in the community, their local clinics, schools, homes, etc. This can help overcome barriers to care due to travel and scheduling, potentially increasing the number of interactions.

This method has its limitations. Interactions over telepresence are not as rich as in-person therapy. Limits from the field of view, resolution, projection of three dimensions onto two, and network latency can decrease the sense of presence of the remote person and hamper their spatial reasoning ^12,13^. The lack of a present person could lead to a reduction in patient motivation. When combined with less clear instructions through telepresence, where seeing what a clinician wants a patient to do can be more challenging than in person, patient compliance might be reduced. If patients are unmotivated and non-compliant, the clinician may not see the motions needed to accurately assess the patient and the patient may not get the personal benefit from the interaction.

One solution to augment the richness of remote interactions is to use Socially Assistive Robots (SAR) ^14^. SARs combine the space of assistive robots, which give aid or support to human users (e.g., traditional rehabilitation robots, wheelchair robots, mobility aides, etc.) and socially interactive robots, whose main task is to interact with people socially. The SAR is therefore designed to create close and effective interaction with human users in order to give assistance, leading to measurable progress in convalescence, rehabilitation, learning, etc. SARs could be coupled with a telepresence system and a computer vision system to improve remote health care delivery by providing a physically present social entity for patients to interact with. The SAR could act as a mediator in the interaction, thus providing a richer experience.

To explore the idea of using a SAR with telepresence, we have designed two generations of systems. Based on our initial prototype (“Flo”, seen in fig. 1) ^15,16^, discussions with clinicians, and the literature, we developed a series of design requirements for our second iteration, “Lil’Flo”. The goal for the Lil’Flo system is to help understand how pediatric patients with motor impairments requiring telerehabilitation, the application of telemedicine for rehabilitation, can be assessed efficiently, cheaply, and accurately. The system enables the examination of two critical components: the telepresence system for communication with the patient and the perception system for understanding the patient’s motor function.

**Figure 1.**
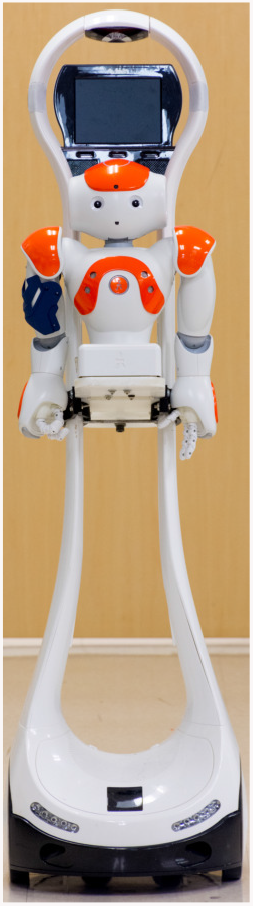
The first version of our social robot on a telepresence system, Flo, composed of a Nao robot attached to a VGo telepresence robot.

Our long-term study goal is to be the first to rigorously compare classical telepresence-based assessment and the use of a social robot as a mediator in telepresence. The ideas of social robots and telepresence are not novel, nor is the study of remote versus physically present agents. The innovation of this project will be to use a social robot as a third agent, physically present with the patient, to mediate telepresence. Many other systems attempt to temporarily replace the therapist with the social robot. Our goal is to extend the reach of the clinician into the community and improve their ability to remotely interact with their patients and accurately track their rehabilitation progress. The social robot would therefore act as a bridge for pathways of communication lost over telepresence (Figure 2). We hope patients will be more motivated and compliant in performing their activities at home and will be able to receive instruction on improving their rehabilitation. This model for interactions has driven the design decisions for Lil’Flo.

**Figure 2.**
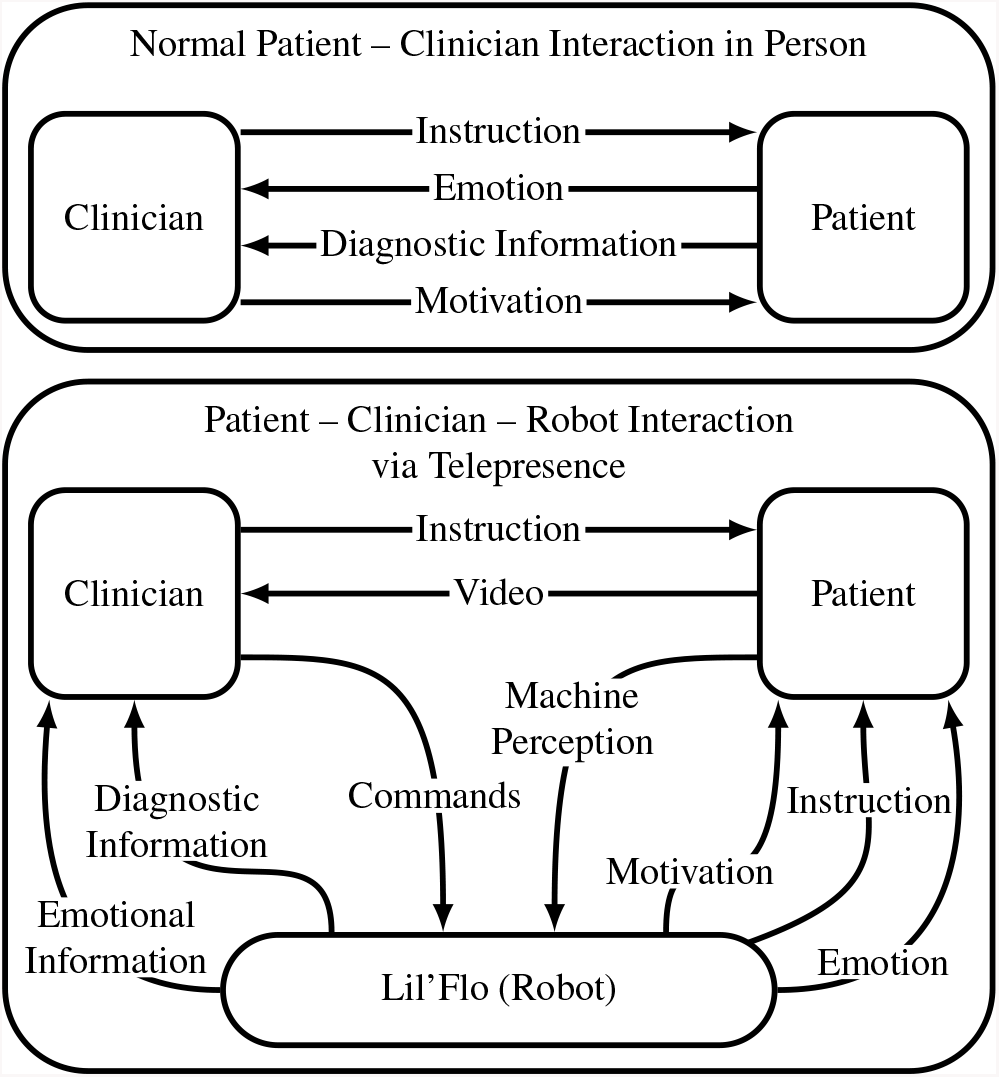
A comparison of our theory of interactions between patients and clinicians in-person and patients, clinicians, and the robot in telepresence interactions. Without the robot in the telepresence interaction, there is a loss of communication channels compared to the in-person interaction.

This paper reviews the related literature, the design requirements for the Lil’Flo robotic system, the design developed, and future directions.

## Related Literature

Rehabilitation robots that are able to gesture, communicate, motivate, comfort, and teach exercises have been tested in pediatric and geriatric interactions in and out of hospital environments ^17–23^. Experiences with these robots have been positive, and the robots have been shown to promote engagement and adherence to prescribed exercise. However, to our knowledge, they have not yet been tested as a tool to augment telemedicine as a third agent to interact with the patient and therapist.

### SARs

A number of social robots have been developed for upper extremity rehabilitation, from which we take inspiration. The Nao-Therapist project initially developed a custom robotic bear named Ursus ^24–26^ and has now moved to a Nao robot ^27^, which is used for upper extremity rehabilitation for pediatric patients. The system uses a Microsoft Kinect sensor to track patients, allowing the robot to autonomously play games with them. It can both demonstrate and correct poses in a pose mirroring game and in a pose sequence recall game. In a longitudinal study of the system, with 13 subjects participating in on average 11.6 sessions of approximately 24 minutes each ^28^, all stakeholders, clinicians, parents, and children, found the system useful and wanted to continue to use it.

RAC CP Fun is another Nao-based robotic platform designed to engage with preschool students who have CP ^18^. The robot can play various games and motivate physical activity. The interactions with the system were designed to build off the motor learning literature, with an emphasis on giving feedback to the patient. The robot interacts by singing songs, changing its position relative to subjects, and providing feedback. Fridin et al. compared outcomes of using the robot between typical children and CP children, finding that the CP group exhibited a higher level of interactions as measured by the child-robot interaction measurement index which relies on eye contact as well as various facial, body, and vocal expressions of emotion ^18,29^.

Another Nao-based system is Zora, which is commercially available. It has been tested on a cohort of children with disabilities and has been shown to improve the quality of care ^30^. However, it was reported that the software required to operate the system was labor intensive for clinicians.

The social robot community has studied questions of robot presence. Fridin et al. demonstrated that when comparing an in-person robot and robot projected onto a screen, pediatric subjects interacted significantly more with the in-person robot ^31^. Bainbridge et al. showed that physical presence is important for trust and motivation, especially for uncomfortable tasks ^32^, and Kiesler et al. showed that subjects co-located with a physical robot were more engaged with it and acted more ideally around it (as measured by following diet advice) when compared to a virtual agent ^33^. Mann et al. demonstrated that subjects were more likely to trust, be engaged with, and follow instructions from a robot giving instructions and asking questions, compared to the same interface on a tablet ^34^.

The idea of socially assistive robots as mediators for interactions has been explored for in-person therapy of children with behavioral disorders such as autism spectrum disorder, where direct human-human interaction can be challenging. The Milo and Kaspar robots are two examples of this with sizable deployments in the clinic ^35–37^.

SARs can easily become complicated systems, it is important to consider how to make systems approachable by being simple and affordable. The CosmoBot system is a good example of how simplifying problems can lead to effective systems. CosmoBot is a small toy-like space robot, integrated into “Cosmo’s Learning Systems”. It has arms with a single shoulder degree of freedom, an actuated mouth, an actuated head, and the ability to drive around. It interacts with patients through a button board, accelerometers placed on the patient, and 3rd party interface devices (e.g., joysticks, buttons). During a 16-week longitudinal study with interactions once a week with four subjects aged 4–10 with CP, it was shown that the system itself was robust and easy to use ^38^. Patients were engaged and excited to play with the robot throughout the length of the study. The system was marketed for a few years by AT KidSystems. Even with its limited number of degrees of freedom in its arms and torso, it was still able to motivate patients to work on their rehabilitation goals. By using the trackers on the patients’ bodies, the robot was able to both interact and collect objective data throughout the study.

Tega is a small smartphone-powered robot designed primarily for education, helping students to develop language skills through interactive storytelling ^19^. It has a design that is supposed to be cute and approachable with five degrees of freedom, allowing it to bob up and down, twist, lean, and look up and down. Tega is inappropriate for most physical and occupational rehabilitation techniques, as it has no limbs. However, it is worth appreciating for its emotional expressiveness, based on principles from animation, and its relatively low cost, using a cellphone for both its face and computational power.

### Telepresence Systems

There have been many telepresence systems developed. The simplest telepresence can be achieved using cellphones, tablets, or computers with a screen, camera, and Internet connection. More advanced systems use a screen on a stick morphology with a screen and camera mounted on a mobile base, allowing the remote operator to drive the telepresence interface. Still more advanced are systems that utilize actuation of the screen based on where the remote operator is looking ^13,39^. Some systems also have appendages which can be moved to show operator intent ^39^ or to achieve a goal.

Dodakian et al. presented a system for stroke patients, using a custom tabletop game system attached to a computer for both individual activities and activities monitored by telepresence ^40^. The system did not use a robot, but by making the games physical, compliance was increased. Using the games with intermediate telepresence provided a way to maintain motivation and provide ongoing assessment.

What we propose is different from these: the telepresence operator is presented in a traditional screen-on-a-stick fashion, while the robot is an independent social entity which can play with subjects and demonstrate actions of interest.

### Objective Assessment

A focus on accessible metrics has been missing from the socially assistive rehabilitation robotics and telerehabilitation literature. In order to make telemedicine feasible from the clinician’s perspective, tools are needed to assess the patient’s function quickly and easily. As shown in the Shriners Hospital Upper Extremity Evaluation (SHUEE) ^41^ and Assisting Hand Assessment (AHA) ^42^, assessment can be done by graders watching recorded video but requires expert graders and is time-consuming, limiting the availability of the test and increasing its cost.

There is a large literature of metrics for upper extremity function that can be evaluated using robots like the MIT-Manus ^43^. These generally rely on the kinematics of subjects, but do not provide the outputs that clinicians expect to work with and require expensive fixed hardware to be used. By extending from this work to use low-cost mobile hardware with computer vision that leverages modern machine learning techniques and working to generate metrics which are intuitive to clinicians, the ability to assess patients could be improved.

## Design Requirements for Lil’Flo, a Socially Assistive Robot for Upper Extremity Motor Assessment and Rehabilitation Via Telepresence

The requirements for the Lil’Flo project are presented below. We discuss our initial prototype and the evolution of the design requirements that lead to those we used in Lil’Flo robotic system.

### Our Original Prototype

The first version of our social robot system was comprised of two individual pieces of hardware: the VGo base and the NAO humanoid (fig. 1). The systems did not interface with each other and were controlled through separate software, which made setup of the robot and system operation difficult, even for the researchers familiar with it. For further experimentation with design, we wanted a more modular system than what the NAO and VGo offered. We found that although the NAO is highly programmable and easy to use, it is hard to modify, and when it breaks, hard to maintain. A computer and cameras were permanently built into the NAO and were unable to be upgraded. The VGo is user-friendly in its default configuration but offers no programmatic interface to extend its capabilities and adding additional hardware is nontrivial ^44^. We found from testing that the NAO platform lacked sufficient sensors to perceive patient interaction and again its cameras are not upgradeable. The NAO has a static face with eyes that can change color; it has been claimed that this, with the pan tilt of the head, is sufficient to convey emotion. But we felt that further facial expressiveness was needed.

The sizing of the system also proved to be difficult, as it is too tall for most pediatric encounters but has a small humanoid for adult encounters. Because of how the structure of the two robots fit together, the NAO’s center of gravity sat in front of the VGo’s center. To counteract this imbalance, an extra weight was added on the back of the VGo, which significantly increased the mass of the system and raised its center of mass, making the system less stable. We used the torso-only version of the NAO, which is no longer sold, and faced challenges with the robot overheating during use. Although newer models have improved on the overheating problem, multiple fellow researchers and collaborators have reported that it can still be a challenge. The NAO/VGo combination was also costly: The current full body NAO is cheaper than when the original prototype was built, but still costs $9,000 USD pre-tax. The VGo robot costs $4,000 USD or $6,000 USD with Verizon 4G LTE with a dock. There is an optional service contract which is around $400 USD depending on the term of the contract. Additional modifications would then be needed to add sufficient cameras and compute power, adding further cost. This type of integration requires significant engineering effort and requires compromises ^44^ which we did not believe were logical to undertake. The cost of components is not necessarily important for research, but by proving that low cost components can be effectively used, an argument can be made that a larger impact can be achieved post technology translation.

The primary takeaways from our initial demonstrations and surveys with the first version of our social robot were that clinicians viewed the robot as a social entity, although they did not find it to be as helpful as we had hoped given its lack of modularity, difficulty in setup, and high cost.

### Design Requirements for Lil’Flo

In developing our new prototype, we decided to rethink some of our prior design assumptions leveraging our new data. Our primary design requirements became that the robot be low-cost, flexible and adaptable to testing, expressive, programmable, and sized for pediatric as well as adult patients. We decided to split our concerns into two sister platforms: one for adult populations (Big’Flo) and one for pediatric populations (Lil’Flo). We are focusing first on the pediatric system, with plans to port the design learnings to a larger adult version with similar capabilities, but a larger size and more adult-oriented aesthetic.

#### Interaction to Support Motor Function

During motor rehabilitation interactions, there are three primary categories of motion that we are seeking to drive: motions that exhibit the range of motion of the patient, motions that allow observation of the kinematics of the subjects’ movement (what does the position vs. time profile of motions look like), and bi-manual motions. It is important that the patient understand what they are being asked to do so that they can attempt the activities. Because different patients will be at different levels of function, the pacing of activities has to be adjustable to the patient on the fly. Ideally, the difficulty of activities should also be adjustable.

Because we are interested in upper extremity rehabilitation and demonstrating movements with a humanoid robot, it is important that the humanoid has arms. At a minimum, the arms need to be able to perform shoulder flexion, shoulder extension, shoulder abduction, shoulder internal/external rotation, and elbow flexion/extension, these are the primary motion planes affected by brachial plexus injuries. For cerebral palsy, wrist supination and even hand manipulation are relevant as well, but our clinical partners have told us that useful testing can be done without them.

With this platform, we plan for all interactions to be seated, which means the humanoid should appear to be seated as well. Although there are some exercises that can be done with the legs while seated, the focus is upper extremity work, and so legs are not critical.

#### Interaction to Increase Engagement

The literature supports the idea that social robots improve engagement and elicit social interactions that keep both pediatric and geriatric patients engaged ^21,28,45–47^. In rehabilitation, motivation is linked to motor function improvement, thus we believe that the use of robots with the ability to interact will be beneficial for patient outcomes. In order to be motivational and build meaningful interactions with subjects, the robot should appear friendly. The humanoid portion of our system is designed to interact with patients socially, and an important component of social communication is facial expression. The design of a face can affect the perceived trustworthiness, likability, and friendliness of a robot ^48^. Speech is also a necessary component for social robot communication.

#### Robot as Peer

The goal for the robot is for the humanoid to interact with the patient as a peer. They should not act as a doctor, therapist, or nurse, but as a friend who is playing games and doing activities. To have mass appeal, the robot should be somewhat ambiguous in its identity: for the system to be approachable to a wide variety of patients, it should not be strongly male or female nor be strongly of one race nor creed. In order to avoid the uncanny valley, the robot should not be overly human.

Ensuring that the robot is interesting, intelligent, sociable, able to communicate, and helpful are important. Engineers therefore must consider what parameters are needed to ensure these features are realized. It is important to highlight that none of this needs to be authentic, the interactions can be a show, as long as they are convincing.

#### Portability

In our new system, we aimed to reduce the weight of the system and lower the center of mass. To allow the system to be tested and used in different locations, it needs to be portable without too much difficulty. It should therefore be liftable by a single person. Within the space which it is deployed, the system should be mobile under its own power. This allows the remote operator to move the robot without assistance, which is important to the feasibility of telepresence to expand access to care. Mobility also improves the sense of presence for the remote operator ^49^.

#### Easy Setup

We received feedback from clinicians that such a robotic system needs to be easy to setup. Although this prototype is not envisioned for unsupervised clinical deployment, ease of setup helps prevent errors by reducing the cognitive load on the operators. Additionally, because the robot is designed to work in medical care situations, it must be easy to clean.

#### Modularity

It is important that the mechanical design is modular at multiple scales. At the individual system level, the humanoid needs to have a head, face, and arms that can be modified and replaced. For the benefit of research, this modularity allows the system to be used as a platform for developing and testing a variety of designs. For example, being able to add extra degrees of freedom to arms, add lighting to the chest, or change the style of face is useful. Modularity is also useful for repairability and upgradeability. At the platform level, to test the efficacy of different systems, namely the humanoid, it is important that they can be removed.

#### Remote Operation

Given the goals of the system, enabling remote operations and telepresence are clearly necessary. However, building a fully secure platform would be beyond the scope of an academic research endeavor. Running from a remote system on a private network is sufficient for research. For a commercial/clinical deployment more security would be necessary. It is important to demonstrate that the telepresence-related technologies work well and reliably, but the interface presented to the remote operator does not need to be aesthetically pleasing at the research stage of development, so long as the lack of beauty does not impair function.

For Lil’Flo’s intended uses, one hour of runtime is required at a minimum, to allow the system to run for the length of a study. For true clinical deployment, a much longer run time would be desired. It is not necessary for the robot motors to be particularly powerful, as the system does not walk or lift anything other than its own arms, relying instead on social interactions to drive rehabilitation goals. It is important that they be sized large enough that they do not overheat when moving the arms.

#### Programmable by Clinicians

Most interactions with this prototype will be programmed and managed by researchers, but both for their benefit and as a proof of concept for clinician-controlled operations, it is important that both programming and operation be as simple and user-friendly as possible. From this, direct constraints were developed for both programming and operation: all operations should be done through a single interface, and interactions should be able to be developed through a graphical interface. Similarly, it is important that the underlying code is easy to understand and leverages existing frameworks to enable future development. The end user should not need to be aware of, competent with, or think about the underlying code during operation.

It is important that users, whether clinicians or researchers, do not need to install any non-standard software. Many robotics systems use Linux-based software. However, Linux is not well-received in healthcare facilities. For general adoption of the system, the technology must be as easy as possible for clinicians to learn to use and require minimal IT overhead.

#### Provide Assessment Information

An important component of ongoing rehabilitation is continuous assessment. Assessment allows clinicians to update the rehabilitation regimen, help give ongoing instruction, and motivate patients. We believe that one of the key use cases for telepresence systems with social robots will be intermittent assessments, while patients complete most of their rehabilitation at home. Doing assessments remotely is possible. The Shriners Hospital Upper Extremity Evaluation (SHUEE) ^41^ and Assisting Hands Assessment (AHA) ^42^ are two examples of assessments that are done using recorded video. Although recorded video can be used to perform assessments, they are time-consuming and must be graded by multiple clinicians to make assessments objective. Clinicians whom we have talked to describe having to watch videos multiple times to accurately assess the patient. At the same time, many assessments fail to be objective or gain a sense of objectivity by significantly reducing the dimensionality of the test. An overview of clinically relevant assessments of upper extremity function are provided by Gilmore et al. ^50^ and Wagner et al. ^51^. It is important that the system we are building provides the infrastructure for exploring remote objective assessment and that the interactions which we design elicit actions that are valuable for assessment.

## The Prototype of Lil’Flo

Figure 3 shows the current generation of the robot with and without the humanoid. Since Lil’Flo is designed to be relevant for upper extremity rehabilitation, it is designed with an anthropomorphic form to allow the system to demonstrate human motion naturally. Because we are targeting pediatric patients, it has proportions similar to those of a child through its arms and torso, with a simple head, reminiscent of a toy, giving emphasis to its face. The robot supports shoulder flexion/extension, shoulder adduction/abduction, internal/external rotation, and elbow flexion/extension. It is built using motors from an XYZ Robotics Bolide robot. The shell around the motors is designed to make the robot more aesthetically pleasing and structurally sound. The robot is designed to be mounted onto a mobile base on which it will appear seated.

**Figure 3.**
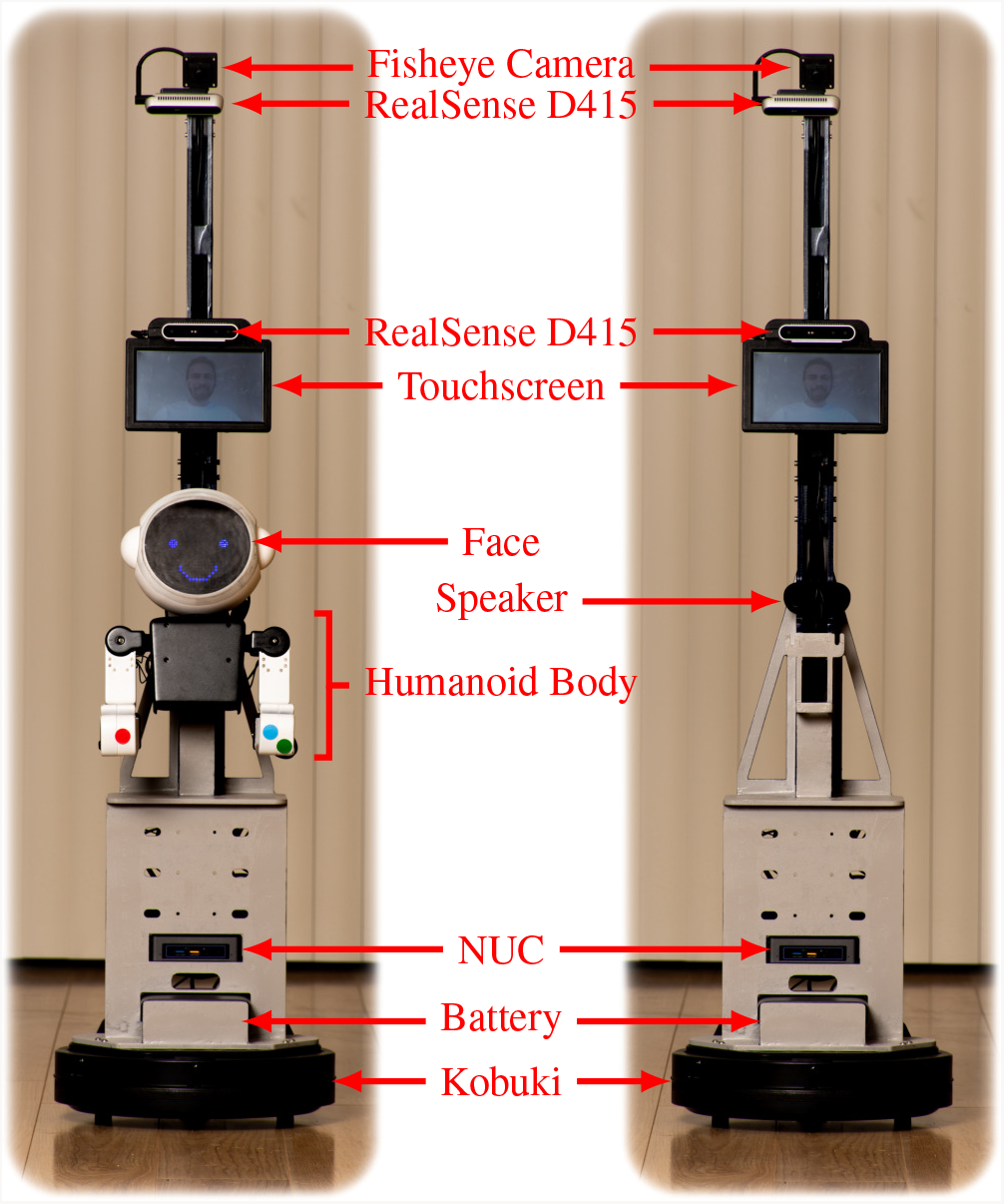
Our second generation socially assistive robot with telepresence, Lil’Flo, constructed using smart servos, a custom frame, and a custom exoskeleton. On the right, the same system, without the humanoid.

The first step in designing the robot was to develop sketches of what the system might look like. Sketches generally fell into the categories of spaceman, toy/doll, and anime (fig. 4). As we continued to refine our ideas, the three major design themes evolved into a spaceman, animal, and child theme (fig. 5). We worked to simplify these ideas, developing primarily with the spaceman theme.

**Figure 4.**
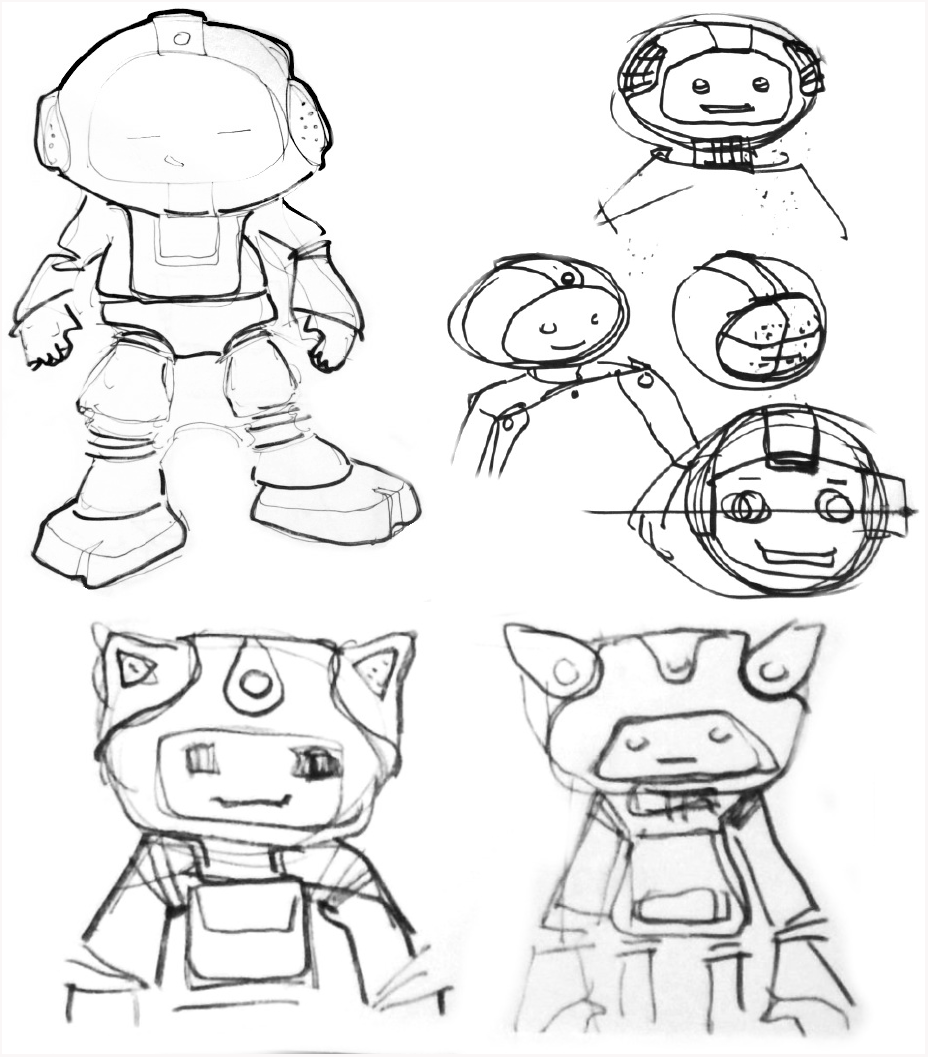
Early concept sketches for the robot showing a spaceman like concept, a toy/doll concept, and an anime like concept.

**Figure 5.**
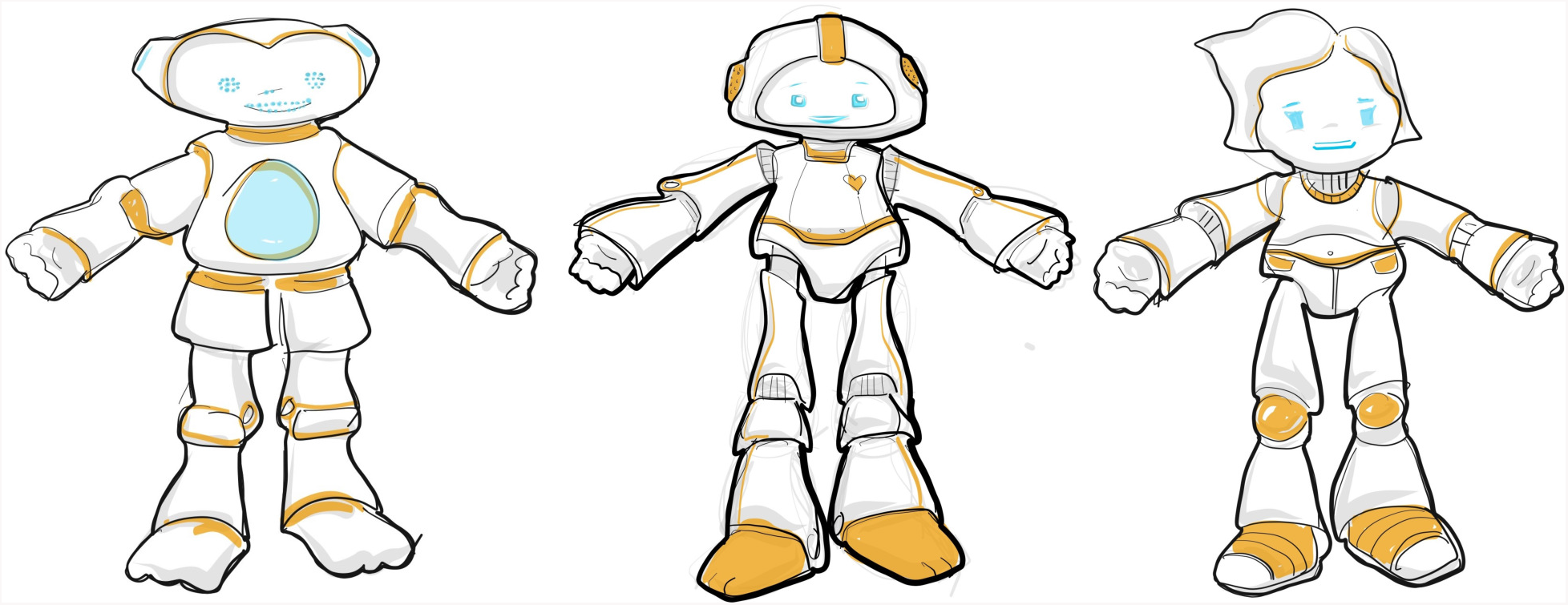
More refined sketches showing an animal, spaceman, and child theme.

We felt that making a generic concept which still had a geometrically interesting form would appeal to the greatest number of people. The color scheme is primarily white and black (fig. 6), providing good contrast along the arms to easily see the components and differentiate body parts. The goal of the neutral color scheme is to give the system a neutral sentiment and allow the face and motions to drive the emotional state, as well as to prevent the robot from seeming to be from any human group.

**Figure 6.**
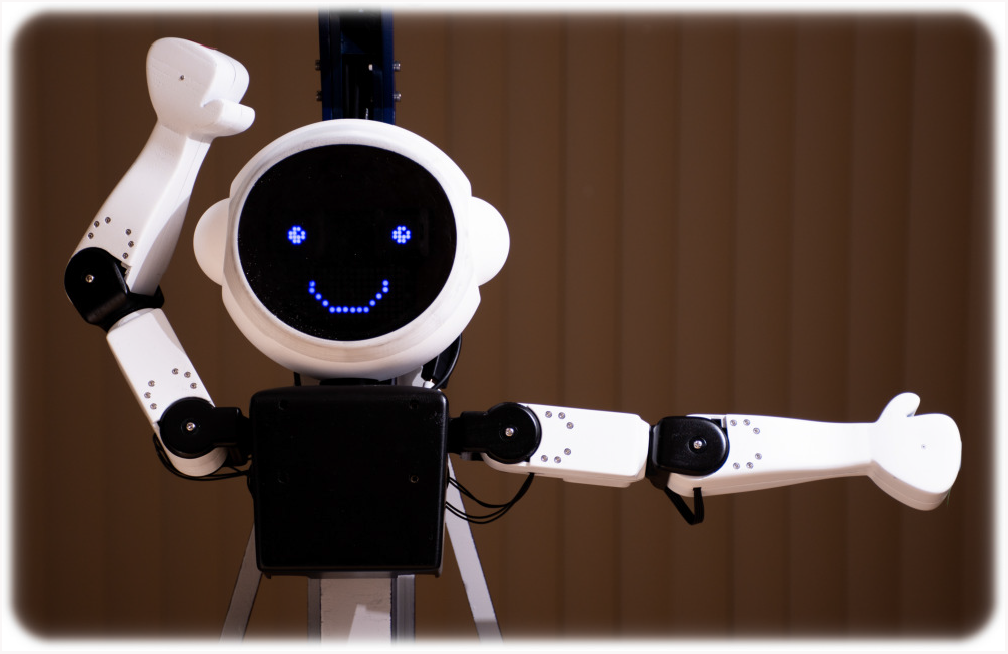
A close up shot of the Lil’Flo humanoid robot. The color scheme uses white arms with black joints and a black chest. The head is white, with a dark screen backed by blue LED lights in a matrix.

### Body and Arms

Lil’Flo uses motors, a control board, and the chest of the XYZ Robotics Bolide robot, a commercially available, affordable, edutainment robotics platform (fig. 6). The Bolide’s motors are serially controlled and can provide digital feedback. However, the Bolide is not appropriate for upper extremity rehab in its native form, lacking appropriate placement of degrees of freedom and having an exposed skeleton. To rectify this, we have developed a custom exoskeleton for Lil’Flo, which has a visually pleasing exterior, and is designed to minimize weight and assembly steps, allowing easy maintenance and experimentation on the robot’s form (fig. 7). The shells have internal structures which give them rigidity while remaining lightweight. The design leverages the rigidity of the motor casings where possible. The motors are placed as high up the kinematic chain as possible, one in the chest, two in the upper arm, and one in the forearm, to improve performance. The motors are fully encased and pinch points are minimized. The hard exterior makes it possible to wipe down the robot, but it is not sealed to fluids or dust.

**Figure 7.**
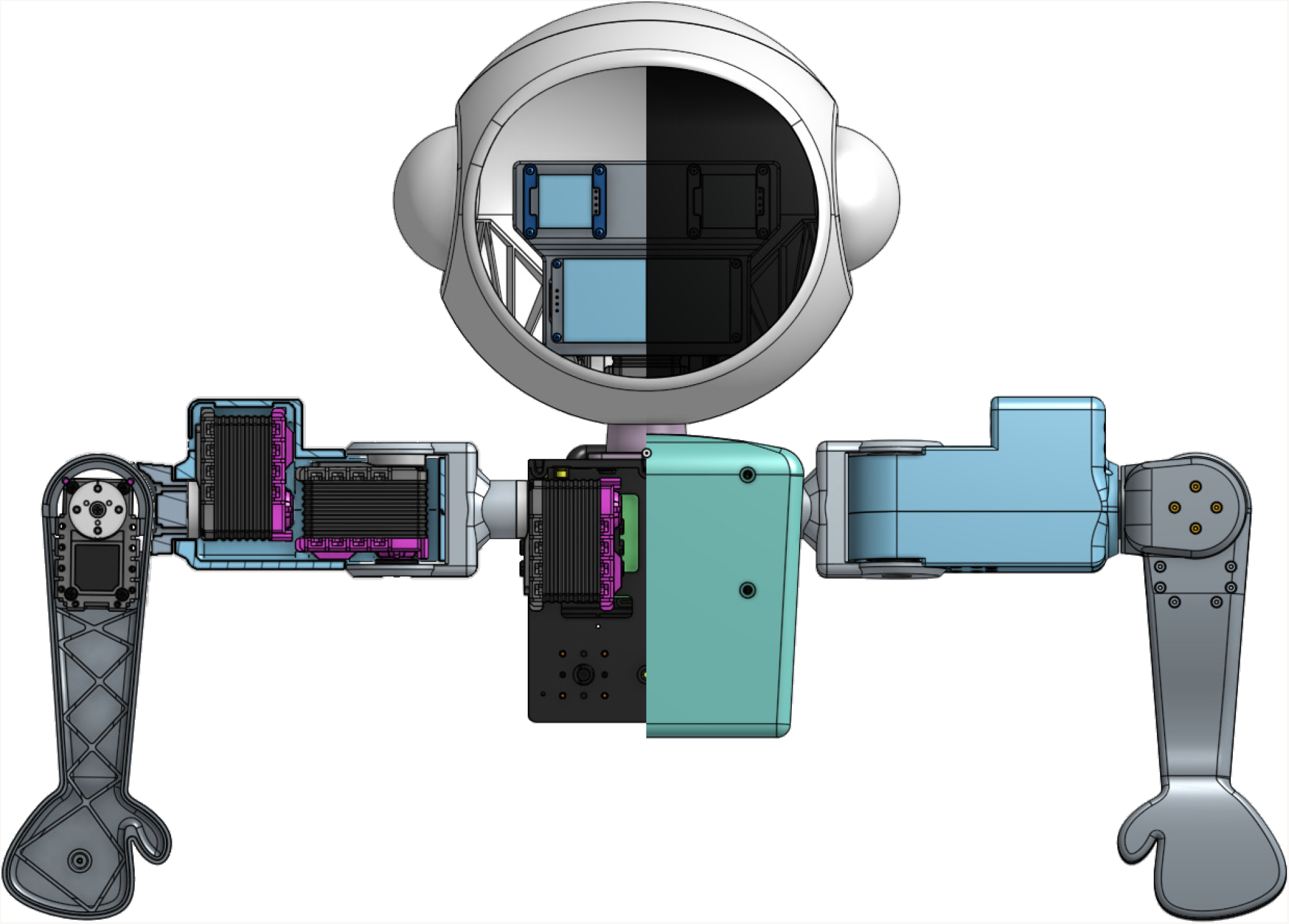
The computer aided design of the humanoid with a cutaway to show the internals of the arms, allowing the orientation of the motors to be seen, and head, allowing the display screens to be seen. The same motors are used throughout the design. Wires are not shown. Fasteners are hidden in the cut out region.

To produce a test ready system, the parts were 3D printed in an ABS like material using fused deposition modeling. The parts were sanded, filled with high build primer, and painted with multiple coats of semi-gloss paint. We felt it important that the parts look production grade to prevent low quality fabrication from being a confounding variable.

The system of motors is controlled by custom software, exposing it to the robot operating system (ROS). The interface works over a serial connection to a microcontroller which performs the real time operations to interpolate the motors over motions. The microcontroller communicates with the motors via a second serial connection. This allows the arm’s movement to be captured, visualized, and controlled in ROS. The Bolide system provides better than hobby grade servos which are digitally controlled with integrated low-level controllers and can provide feedback to the higher level system. These motors were selected because they are cheap, serially controllable servos that can generate enough torque for our system (stall torque of 25 kg-cm), sufficient to wave the arms around.

### Head and Face

Our first physical prototype for the head can be seen in fig. 8. The prototype highlighted the importance of having a screen on the face which shows only the eyes and mouth while hiding the internal mechanics and gave direction to the correct proportions for the head. During prototyping, an informal straw poll between a head with and without ears showed that the ears were preferred, as they gave a sense that the robot could hear.

**Figure 8.**
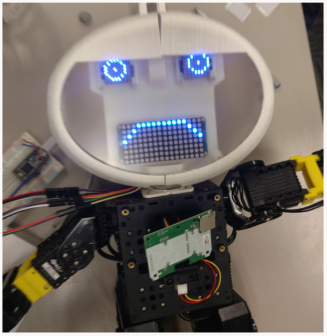
The first physical prototype of the head for the robot. The head is too wide for the body and the clear screen exposes all the internal components.

The current head is broken into three major components, a front shell with a translucent urethane panel, the back, and an inner skeleton which holds the LED matrices (fig. 7). The front and back sections connect with a seam that follows along the back of the ears and above the top of the head, making a clean line which could be interpreted as a hairline. The head is 3D printed out of white material and sanded.

Because the face is often the center of attention for human-human interactions, we want the flexibility to alter the facial expression on the fly. Initially, LCD screens were explored to create the face, but it was challenging to get the geometry of the head and face to work with a single large screen while being affordable and having a bright screen. LED matrices solved these challenges. The primary compromises with using LED matrices are that they are single color and have low resolution. The other option for making a digital face is to use a projection based system, as Quori uses ^52^. The projection system can handle complex geometries and provide good brightness. However, fitting a projector and reflector in a small platform is challenging and costly.

We ultimately designed a head with a dark translucent face and variable brightness LED dot matrices behind the surface (fig. 6). Three LED dot matrices, one for each eye and one for the mouth, are controlled from a Teensy microcontroller which provides power to the matrices and communicates with them over serial. The microcontroller then communicates with the remainder of the system and receives power via a USB connection. The LED matrices allow us to alter the expressions of the robot while keeping the face simple from an aesthetic and maintenance perspective.

To achieve the dark face, a translucent black urethane screen is molded into a thermoplastic 3D printed shell of the head. Molding is used rather than other techniques to allow the screen to have a geometrically interesting shape, with a curved forehead, and to allow the piece to seamlessly exist within the rest of the head. This is done by making a positive of the face and screen together and pouring a mold to match it (fig. 9). This yields a mold which fits the front of the face and defines the shape of the screen (fig. 9). The 3D printed face is then placed into the mold and a clear urethane which has been mixed with a black colorant is added (fig. 9).

By using a dark face, we provide good contrast to the head with clean lines, highlighting the face as a point of attention. The dark face also allows us to hide the internal structure of the head while allowing good transmission of light from the internal LEDs, leading to good facial feature visibility.

**Figure 9.**
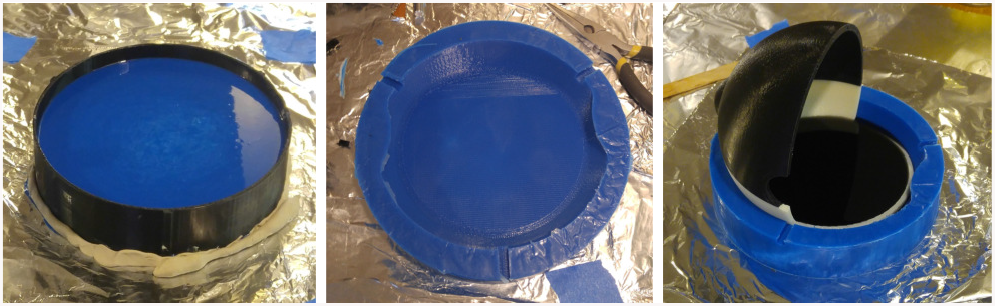
On the left, making a mold to make the face screen. In the middle, the completed mold for the face screen, which is used to mold the translucent front screen. On the right, the translucent front screen being molded into a prototype 3D printed model of the head. The material being used is a clear urethane with black colorant added.

Finally, a series of patterns for the LED matrices had to be designed. Eye and mouth proposals were developed, taking inspiration from emoticons and general facial expressions. These were distilled down to mouth/eye combinations which we felt would be most relevant to our use cases. Some of the eye sets are directional (i.e. able to look left, right, up left, down, center, etc.), others only have a single direction.

### Base

The base of the robot is built with an off-the-shelf Kobuki mobile robot platform with a custom set of risers to hold the computer, humanoid, cameras, and screen. The Kobuki was selected because it is affordable and integrates well with the Robot Operating System (ROS). It also provides mounting points to build custom systems off. The structure of the remainder of the frame is constructed of MDF, which was laser cut, glued, filled, smoothed, and painted. The paint uses a primarily gray color in satin with blue accents. There are three distinct areas of the base: The first section is the bottom, which attaches to the Kobuki, houses the computer, a USB switch, a battery, and excess wiring (Figure 10). The bottom area also has mounting holes that were used to explore camera placement. The middle area mounts permanently to the base and holds the humanoid. The humanoid hangs from this section, held by gravity and friction, allowing it to be easily removed. The middle area also houses the screen with one of the cameras. The screen is held within a custom 3D printed casing that also provides a mount for a camera. The top section is mounted to the middle section using screws. This was added to the design based on feedback from clinical partners to allow a better field of view for cameras by getting them higher up. The top section holds two cameras.

**Figure 10.**
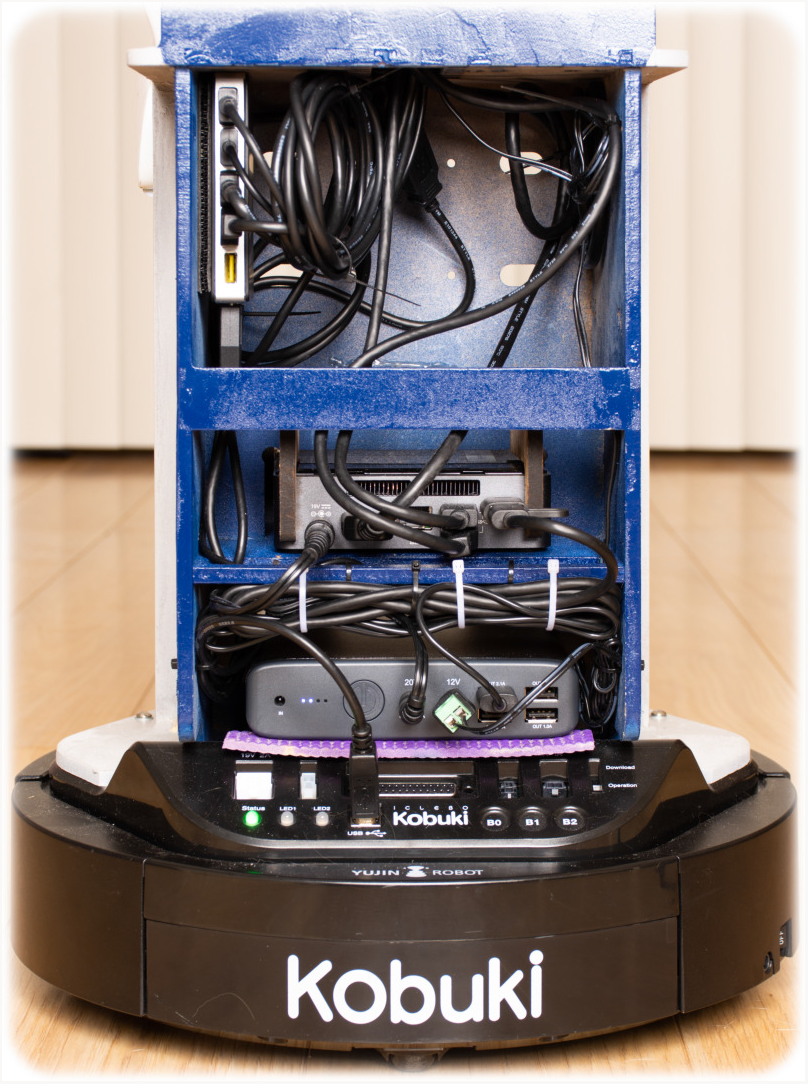
The inside of the lower base where the battery, computer, and wiring are housed.

The base height was selected after some trial and error to make a stable system. The middle of the face of the robot is around 72 centimeters off the ground, the middle of the screen is about 90 centimeters off the ground, and the top of the robot is about 132 centimeters off the ground. For interacting with people who are sitting, this produces a system which is low to the ground, but higher than a Nao robot on the ground which has been used in multiple other SAR focused rehabilitation studies. When sitting, we have found these heights to be on the short side of comfortable for an adult. We did not want the humanoid to tower over a subject and because the system is designed for children, being on the shorter side may actually be an advantage.

### System Architecture

The entire system runs on an Intel NUC NUC7i5BNK with a solid state drive and 16 gigabytes of RAM. Connected to the NUC are a pair of Intel RealSense D415 cameras and a USB 3.0 powered hub (fig. 11). The hub is connected to a 180 degree fisheye camera by ELP, a small speaker which uses USB for both power and input, a touchscreen panel, the robot’s face, the robot’s body, and the Kobuki base. The NUC is also connected by HDMI to the touchscreen panel. The screen is a 7 inch 800×480 pixel TFT screen with a resistive touch overlay. The screen is driven by a driver chip, which provides the HDMI and USB interfaces. The NUC has a dual microphone array built into its front panel, which is exposed through the robot base.

**Figure 11.**
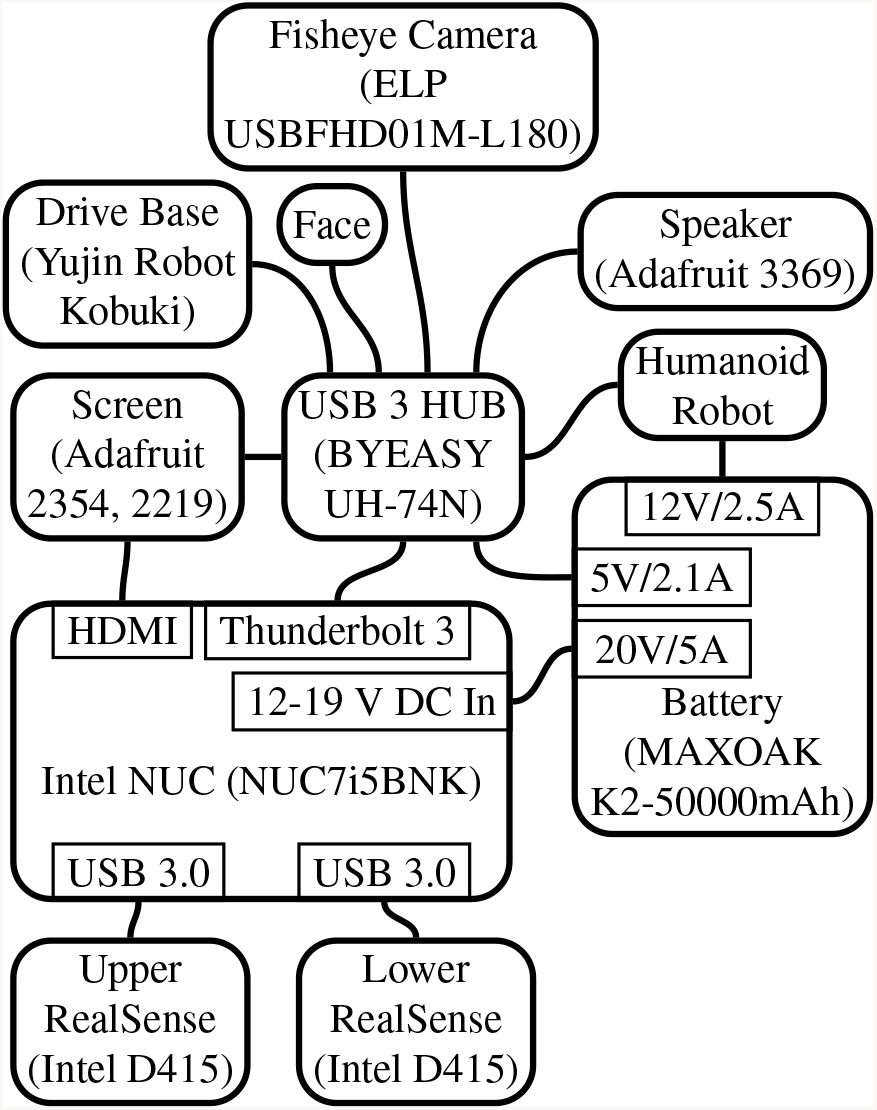
The system’s primary components. The NUC has two USB 3 ports on its back panel as well as a Thunderbolt/USB 3 type-c connector, a HDMI port, and a power port. The RealSense cameras are each connected to an independent USB port to guarantee that they have sufficient available bandwidth. A powered USB hub is connected to the type-c port. The USB hub, NUC, and motors within the humanoid are all powered by a MAXOAK 185 Wh/50000 mAh battery. The screen uses a USB connection for both power and to provide touch input to the computer. The Kobuki base is only connected via USB. It maintains an independent power system. The microphone is built into the NUC and so not shown. The speaker, face, and fisheye camera all use USB for both power and data.

Audio is captured by the microphone array built into the NUC. Because the NUC is placed about half-way up the base, this provides reasonably good sound quality. Audio for Lil’Flo’s voice and the remote operator are provided through a speaker behind Lil’Flo’s head. The remote operator is shown on the TFT display.

*Cameras* Video is captured by two Intel RealSense cameras and a fish-eyed camera. The Intel RealSense D415 cameras are used because they give reasonably good depth results, when compared to similar cameras, but do most of the processing on board. They are low-cost, low-power consumption, light-weight, compact, and have a minimum z-range less than 0.3 m. Because the RealSense cameras use true stereo, they can tolerate uses in environments with multiple other cameras and with strong infrared light, such as near windows. The D415 places all of its imagers on a single rigid plane, allowing them to stay calibrated to each other. The sensors use a rolling shutter, but for low speed human tracking, that is sufficient. Because of the relatively narrow field of view for each sensor, they have a small angle between each pixel, giving fine resolution, which is useful for pose tracking. They also use RGB+IR sensors for their depth stereo pair, allowing them to more easily find features in color rich environments.

A challenge of building the system was getting cameras placed in such a way that they could see the subject at a variety of distances. In order to tolerate both full range of motion activities and activities touching the robot, across a range of ages, the field of view needed to be broader than what one camera could provide. We initially placed one camera above the screen and one lower on the base to serve the two different fields of view. However, looking at peoples hands from below was awkward for analysis and even more awkward for people interacting with the robot (fig. 12, first image). We explored placing two cameras above the screen, but that did not give good coverage of the space where people would be touching the robot, both near their hands and head (fig. 12, second image). During a demonstration, our clinical partners suggested that we needed a larger field of view than we might expect to accommodate the mobility of children, even when seated. They recommended we move at least one camera higher up or behind the robot. To accommodate this, we designed an additional mast for the base which screws on. The mast adds 29 centimeters of camera height from the top of the screen. At the same time, testing with the RealSense cameras had shown them to be good for interactions with a single person, but did not provide sufficient situational awareness for driving or when additional people to the side of the robot tried to interact. So we added a fisheye webcam quality camera from ELP. We examined placing all cameras at the top of the mast, which allows for easy image stitching, but does not provide an expanding field of view at distance, nor does a good job of covering the head area for tall subjects close to the robot (Figure 12, third image). We settled on placing the fisheye camera at the top of the mast with one RealSense camera and the other RealSense above the screen (Figure 12, fourth image). This configuration gives good coverage of the arms and hands for interactions close to the robot from the RealSense on the mast, good coverage of subjects at distance for full arm range of motion activities, with the ability to back up to scale the field of view to the subject, and good general visibility through the fisheye camera. The downside of this configuration is that there is a blind spot between interactions which are near and far away and the images would be challenging to stitch together, so must either be treated separately or brought together as point clouds. Because of this, we chose not to synchronize the cameras, which is a feature available in the D415. Synchronizing the cameras can place more load on the processor and USB bus, but is desirable if images are being stitched together.

**Figure 12.**
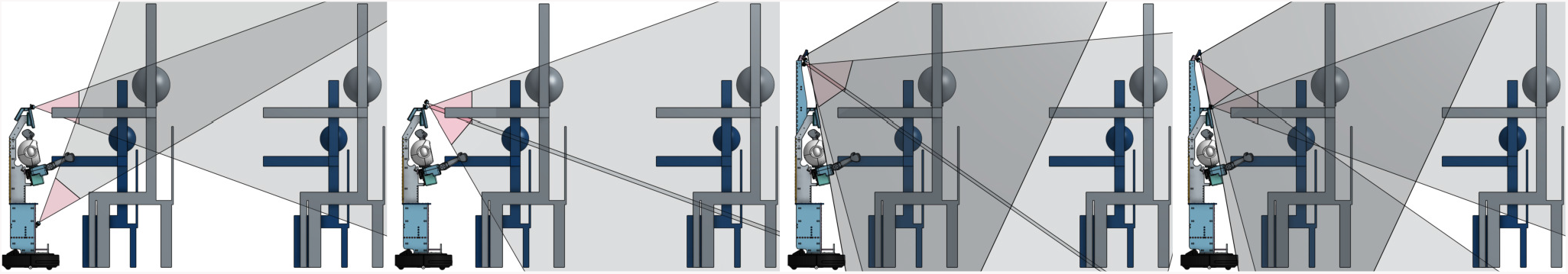
Four different configurations for the cameras. The gray areas show the field of view. In all configurations, two Intel RealSense D415 RGB+D cameras are included. In the last two configurations, an ELP 180 degree fisheye camera is also shown. The depth cameras have a red area shown, which shows an area where depth cannot be recovered. There are two sizes of example subjects shown, an approximate small six year old and an approximate large eleven year old ^53^.

### Software

Figure 13 describes the software stack. The entire software stack is built to interface with the Robot Operating System (ROS) ^54^. ROS was selected because it has become the de facto standard for robot integration. By using ROS, the development of the system can take advantage of a significant amount of work which has been done by others and help contribute components to the community. ROS also provides an opinionated and easy to use system for integrating complex systems. The software stack which we have developed is separated into a series of different components. This infrastructure also makes it relatively easy to operate in a simulation mode for development and testing. The robot currently runs on Ubuntu 16 with ROS Kinetic and the development machines on Ubuntu 18 with ROS Melodic.

**Figure 13.**
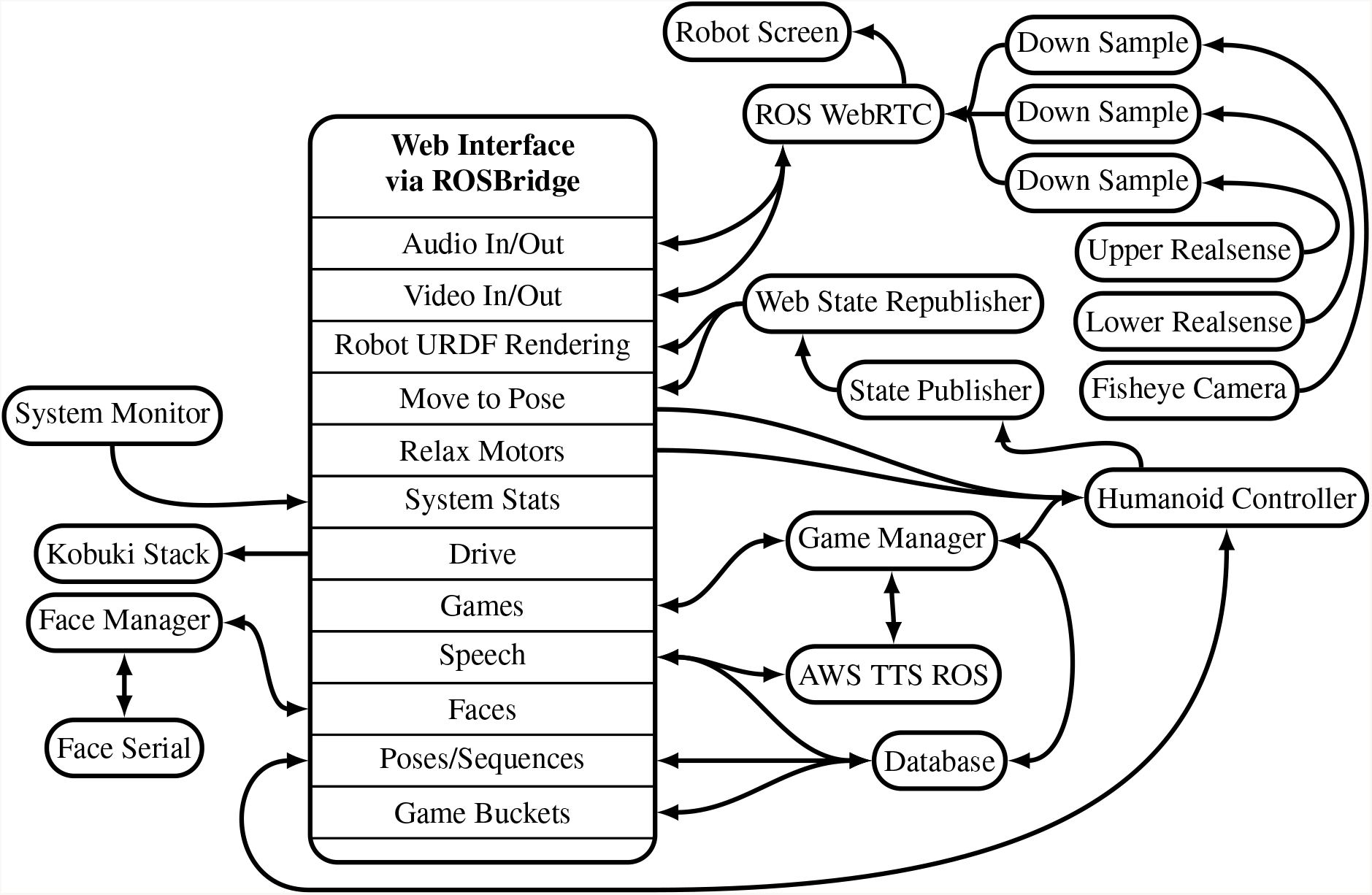
The nodes and their connectivity running on the robot. The entire stack is extremely complex, with many messages passing between nodes, so here we are showing a simplified “logical connection map” and omitting the connections to the recording system, which records from nearly every node.

Our system primarily uses a plays and scripts style of operating under traded control ^55^, allowing the operator to easily manage the complexity of the system. This comes with somewhat limited control from the operator, which is mitigated through options to control the system more directly as explained below. Although we have seen a lot of progress towards it, autonomy remains out of reach for reliable human robot interaction, so using human-in-the-loop systems can allow complex applications like ours to be feasible ^55^. The clinician can provide perception and reasoning that a robot cannot yet provide.

The face is controlled by a pair of ROS nodes, one for handling the serial interface with the Teensy microcontroller mounted in the head and one for handling the face state and commands. The control node exposes a series of services which allow the face, brightness, and eye direction to be set, as well the available faces to be requested. When there is a change made to the active face, the control node publishes the new state. The available faces and eyes are stored in a JSON file. When the system is in a simulation mode, the control node runs alone.

There is a single node to interface with the humanoid. It communicates over serial, with the microcontroller located within the humanoid, based on the XYZ robotics software. Individual poses are loaded from the computer onto the microcontroller to build a dictionary of available poses, and then sequences are loaded as matched pairs of pose IDs and times. The ROS node exposes an action server which receives a series of joint targets which define either a single pose to move to or a series of poses to move through. Sequences of poses are interpreted to create linear motion in joint space between successive targets on each joint. So if the left arm is commanded to a position at 1 seconds and 5 seconds and the right arm is commanded to a position at 2, 3, and 5 seconds, the left arm will interpolate from its position at start to the first position at 1 second and from there to the second position at 5 seconds, ignoring the joints on the right arm, which will be interpolating independently. By using repeated targets, this gives complete flexibility of motion. For example, if the left arm and right arm should move together to 1 seconds, the right arm should move to another pose by 3 seconds while the left arm stays still, and then both arms should move together to poses at 5 seconds, then right arm would be given targets for its first pose at time 1 seconds, second pose at 3 seconds, and third pose at 5 seconds while the left arm would be told to go to its first pose at 1 seconds, its first pose again at 3 seconds, and its second pose at 5 seconds.

There are a series of software checks which ensure that the commands are delivered successfully to the microcontroller. Once the sequence is successfully sent, the action server provides feedback to the calling system on motion progress. In addition, the robot provides frequent status updates to the controller on the current position of the arms, which the control node publishes. These messages are used to maintain the state of the robot and to allow the poses to be saved.

There is a core package which handles launching the entire system, maintaining a database for the robot, and running games on the robot. The database stores poses, sequences of poses, utterances (things the robot can say), and game buckets in a SQLite architecture. JSON is used to expand the standard SQLite data types to handle arrays of various types. The poses are stored with an id, description, and joint name / angle pairs. The poses are side agnostic and can be selected for either side by consuming applications. The pose sequences are stored with an id, description, sequence of times, targeted arms, and pose ids (references to the poses table). Access to the database is exposed through a series of services which perform error checking on row creations and modifications. The database can be searched or directly indexed and modifications can be made through direct index.

#### Image and Audio Capture

Data is read from the RealSense cameras using the stock ROS RealSense packages, which generates output seen in fig. 14. We use the D415’s onboard vision processor to generate depth data and only publish the depth and color feeds. This saves bandwidth and processing power. The depth feeds run at 1280×720 pixels, which is the value recommended for pose extraction by Intel. The color feeds also run at 1280×720 pixels, which is less than their maximum 1920×1080, a compromise to save processor overhead and storage space. All feeds run at 30 frames per second. We do not align the depth to the color or generate point clouds at the time of capture, to minimize processing. These steps are completed as part of a post processing pipeline.

**Figure 14.**
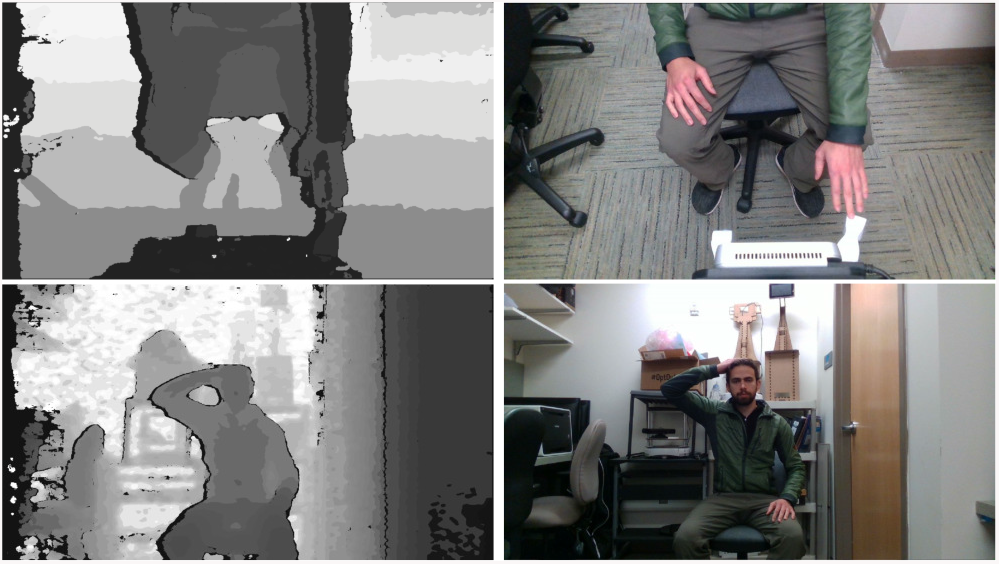
Example output from the RealSense D415. On the first row, the upper camera looking down on a hand touching the robots hand. On the bottom a person touching their head, captured by the lower RealSense. On the left are visualizations of the depth data and on the right the color images. The demonstration subject is a member of the research team.

The fisheye camera video is captured using the video stream cv package, which was the only available package for webcam/usbcam capture that did not require significant modification to other software running on the computer and which ran at the full 30 frames per second. The fisheye camera runs at 640×480 pixels. It is capable of running at up to 1280×1024, but the quality of the imaging optics are not sufficient to justify the compute and storage costs of running at that higher image size.

There is an image processing node for each color video feed which downsamples and republishes the feed at a lower resolution, 300×168 pixels for the D415s and 320×240 pixels for the fisheye camera. All cameras are also throttled to 15fps in the down sampling pipeline. This allows the feeds to be sent to the web interface at a lower resolution, saving encoding processing and bandwidth. The full resolution and frame rate feeds are saved in bag files for future processing.

There is a node that runs to capture audio. This node simply captures audio from the microphones and publishes it within the system.

#### Speech Synthesis

There is also a speech synthesizing node. The robot’s voice is synthesized using Amazon Polly through the Amazon Web Service (AWS) Robotics TTS-ROS package. Other methods of synthesizing voice were considered as well, including solutions from Microsoft Azure, Google Cloud Platform, Nuance, Voicery, and Acapela. We chose the AWS solution because it is low cost, with zero startup costs, flexible, produces high quality utterances, has a rich API, and had the extra ease of a preexisting ROS integration.

Settling on using the AWS Polly system, we chose the Salli voice. Initially we used the Ivy voice, but with time, found it to be overly childlike, somewhat annoying, and difficult to understand for some team members. The Salli voice is warm and soothing, easy to understand, and weakly aged. In order to allow the system to perform in challenging network situations, we modified the AWS ROS TTS node to incorporate caching, caching audio files to the disk with a separate SQLite database to maintain references.

#### Robot Exercise Games

There are currently two exercise game types implemented, a Simon Says game type and a target touch game. In Simon Says, the robot first reads out a set of instructions, which inform subjects that they should do what the robot says and does if it says Simon Says. It then demonstrates steps for the subject to perform, giving them instructions on what to do. For example, “Simon Says touch the top of your head with your right hand”. Thirty percent of the actions are repeated without a “Simon Says” command. The order of the steps is randomized. In the target touch game, the robot moves its hands to different poses asking subjects to touch the colored dot on the robot’s hand with a specific subject hand ten times.

The custom game runner manages the operation of games. Games exist in two parts: a game type and a game bucket of steps. Game steps can be either a pose to move the left arm to, a pose to move the right arm to, a pose to move both arms to, or a sequence of poses. In all cases, the target is a reference to a database object, either a pose or a sequence. Each step also has text associated with the step and an optional time parameter which can be used to set the length of time the robot should use to reach a pose or modify all the times in a sequence to complete the sequence in the specified time. The games are abstractions in code which are given a bucket of step definitions and return a sequence of actions with the fully expanded pose or sequence as a series of joint targets with times and a speech component. This allows additional games to be added as simple middleware with a clear API and no need to think of the underlying robot mechanics.

To begin a game, any other node can send the game runner a message to load a game of either Simon Says or target touch, with a bucket of steps. The game runner processes the game and waits for a command to begin. The game runner publishes two topics, one for feedback and one with command options and listens for commands. The commands can include: start to begin the game, next to go to the next step in the game, repeat to repeat the prior step, congratulate which will make the robot say a congratulatory sentence randomly selected from a bucket of options, try again which will make the robot say an utterance from a bucket of options that communicate that the subject should try again and then run the prior step, quit game which will exit the currently running game, and finish game which exits the game upon completion.

#### Remote Operation

The entire system is designed to be controlled remotely. This is done through a custom web interface (fig. 15) written in typescript, using React. The web interface uses the rosbridge suite on the robot to gain access to actions, services, and topics provided by the rest of the stack, the tf2 web republisher to get access to the transformations which define the robot, ros3djs to show a rendering of the robot, and WebRTC ROS to send video from the robot to the web and back, all from the Robot Web Tools project ^56^.

**Figure 15.**
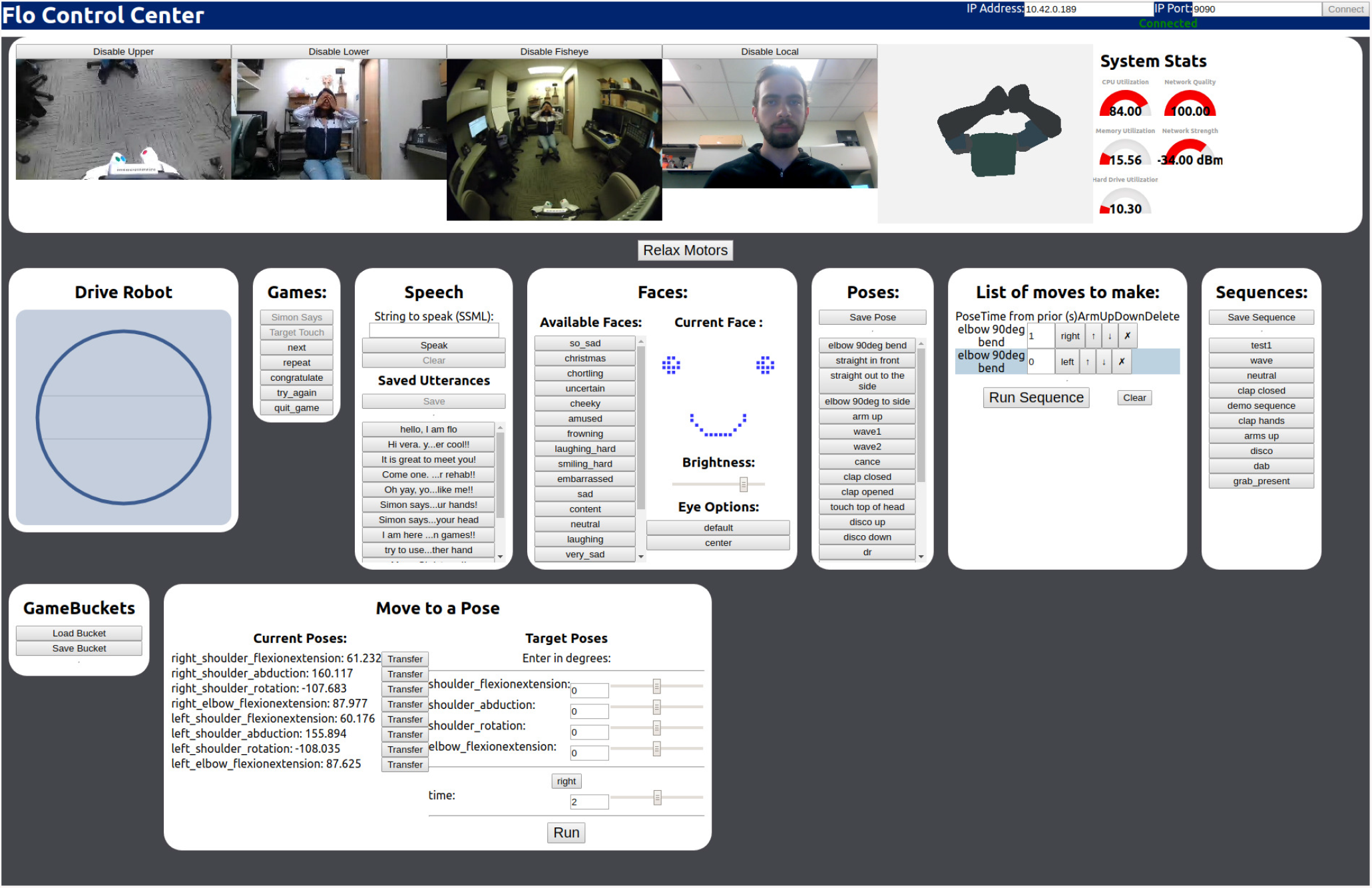
The web interface for controlling the robot. On top are the two video feeds from the RealSense cameras, the feed from the fisheye camera, and the feed from the local camera. There is a rendering of the robot in its current state and system stats for the robot computer showing the CPU utilization, memory utilization, hard drive utilization, network quality, and network signal strength. Below are modules for driving the robot, running games, making the robot speak, controlling the robot’s face, saving and running poses and sequences, creating game buckets, and manually moving the robot. Not shown: a number of the buttons for saving and loading provide pop-ups with further options to achieve functionality. The operator and demonstration subject are both members of the research team.

On the top of the web interface is a bar to set connection parameters which in most situations will correctly auto populate and a button to connect. There is then an area which shows outputs to the operator: the three video feeds from the robot, the video feed of the operator which is being sent to the robot, the rendering of the robot’s pose, and system stats for the robot’s computer. This allows the operator to see where the robot is. The downward facing camera provides a good view of ground obstacles and the fisheye camera allows a wide field for situational awareness. The rendering of the robot state is done from sensors in the motors, not from the planned state, so it provides quality feedback to the operator. The system stats are important, especially the network stats, for working over telepresence, where driving out of network range can leave the robot stranded. All the video is transferred using WebRTC and the video of the operator is captured using the browser based getUserMedia stack.

There is then an area of controls for the robot. There is a button to relax the motors, which is useful for programming poses by moving the robot in person. The drive console presents a circle for the different directions the robot can drive, the user clicks with their mouse to drive the robot. The games module connects to the game manager to select games and run them by dynamically displaying the available game commands and sending them back to the server. A speech module allows the user to arbitrarily type speech for the robot to say, save speech which the robot has said, and select prewritten text. The faces module allows selection of the stored faces, shows the currently displayed face, allows the brightness of the LEDs to be set, and allows the direction of the eyes to be set. The poses, list of moves to make, and sequences modules all work together to define new poses based on the current robot’s pose and define, edit, and run sequences as a list of poses with targeted completion times and arms. The game buckets module allows the user to load game buckets for editing or create new game buckets. Finally, the move to a pose module allows the remote user to manually move the robot through software, this is useful for training new poses in simulation.

To display the video from the operator on the robot’s screen, a small custom program was built which displays the remote video topic using OpenCV. The display runs full screen and on top of all other windows, ensuring that the experience for the patient is not disturbed by pop ups or notifications.

ROS is used to launch the system through a cascade of launch files. There are two entry points to the launch chain, one to run in siumulation and one to run on the robot. The simulator launch file simply launches the standard stack with some added parameters. In practice, the system is started by running a script on the robot (generally by ssh) that starts tmux, starts a roscore, launches the system using rosmon, starts the web server, and starts convenience panes to allow setting the robot’s volume, monitoring detailed performance, and setting the default audio device. At that point, a user on the network need only point their browser to the appropriate IP address, and they are ready to operate the robot. For lower level administrative access the user can ssh into the robot and view the status of the internal system directly.

### Data Collection

All relevant data is recorded in rosbags, this includes all the video feeds and their respective information, all the commands to run the robot which operate via ROS topics, the mobile base information, the robot pose information, and logging output. The rosbags are split every one minute to keep them small and prevent loss of data from corruption at system startup and shutdown. We initially attempted to record the image streams in their raw format, which leads to large bag files, approximately 18 GB per minute. If the image streams are recorded in their compressed format, the bag files are around 3.1 GB per minute. If further lz4 compression is used during recording, the bag file size is about 2 GB, however compressing the depth images costs significant processing power, leading to dropped frames. If the video files are recorded compressed, but the depth is left uncompressed, the size is about 4.3 GB, even with lz4 compression. Our current system has approximately 185 GB that can be devoted to bag storage, meaning that we can store approximately 40 minutes of data using compressed video streams and raw depth streams, with lz4 compression. We have attempted to add storage using an external USB 3 drive, with the plan of recording raw images to decrease processor load and preserve all data, but we found that it could not handle the bandwidth of data and dropped frames.

### Interaction

Using all the tools outlined above, we can construct how an envisioned interaction proceeds. In order to use the robot, the robot must be turned on. A remote operator on the same network (potentially via a VPN) can then login to the robot, start the system software and connect through their web browser. The operator can then navigate the robot to the patient and introduce themselves and Lil’Flo. The operator can ask the patient if they want to play a game with Lil’Flo, select a game type, and select a game bucket relevant to the patient. As the patient plays the game with Lil’Flo, the operator has direct control of the progress of the game, it is a single button click to go to the next step, repeat the prior step, and/or give feedback. The operator retains the flexibility to type in custom text for the robot to speak and, at any time, send the robot to a different pose to probe the patient further. As the patient struggles or does well, the operator can change the face on the robot to express empathy for their struggle and joy for their success.

As interactions progress over time, the operator may choose to modify the game buckets available for different patients. All the while the system will be collecting and logging data on the interactions and how the patients are moving.

## Design Evaluation

As we have progressed through the design process, we have evaluated the success of our designs against our design requirements.

### Upper Arm Workspace Evaluation

To understand the viability of the new arm design, the range of motion of the robot’ joints were measured using the encoder built into the motors. The arm motions were compared to standard human joint range of motions for the shoulder and elbow. Measurements from the constructed robot of range of motion show shoulder flexion: 3.6 radians, shoulder extension: 3.9 radians, shoulder abduction: 3.0 radians, shoulder adduction: 0.1 radians, shoulder internal rotation: 2.4 radians, shoulder external rotation: 3.2 radians, elbow flexion: 1.5 radians, elbow extension: 0.68 radians. The limits are imposed by both physical contact between components and length limits on the wires connecting the motors. The range of motion of the arms provide coverage of human range of motion except for shoulder adduction and elbow flexion. Elbow supination and pronation are not present in the design, nor are any wrist motions. The shoulder internal/external rotation is done near the elbow joint, instead of near the shoulder. We believe that this is sufficient to meet the mechanical requirements for creating interactions to support motor function.

### Face Expression Evaluation

To test the head alone, we isolated it from the body and presented it to 10 subjects. The robot acted in a static mode, dynamic mode, and an iterating mode. Subjects first experienced the head/face in both the static and dynamic modes, while being asked questions about it, both open-ended and on a Likert scale. The subjects were then shown all the faces on the robot and gave the first thoughts that they had. The results are reported fully in our prior publication ^57^. The questions comparing the various face designs and some of the open-ended statements were particularly interesting and helped guide the final design of the robot.

Based on the results we culled the set of faces which we had developed down to a final set, shown in fig. 16 which clearly showed an emotion. The data suggested that a limited face, like the ones shown, can convey gross emotions clearly.

From the open-ended questions during the interview, a number of important points were made. The electronics on our system suffered from interference, which caused some flickering in the eyes and face. The subtle flickering was enough that a number of subjects mentioned it as being detrimental to their interaction with the robot, leading to a redesign of the power system to use higher quality wires in twisted data and power pairs with quality connectors going to the microcontroller and matrices.

**Figure 16.**
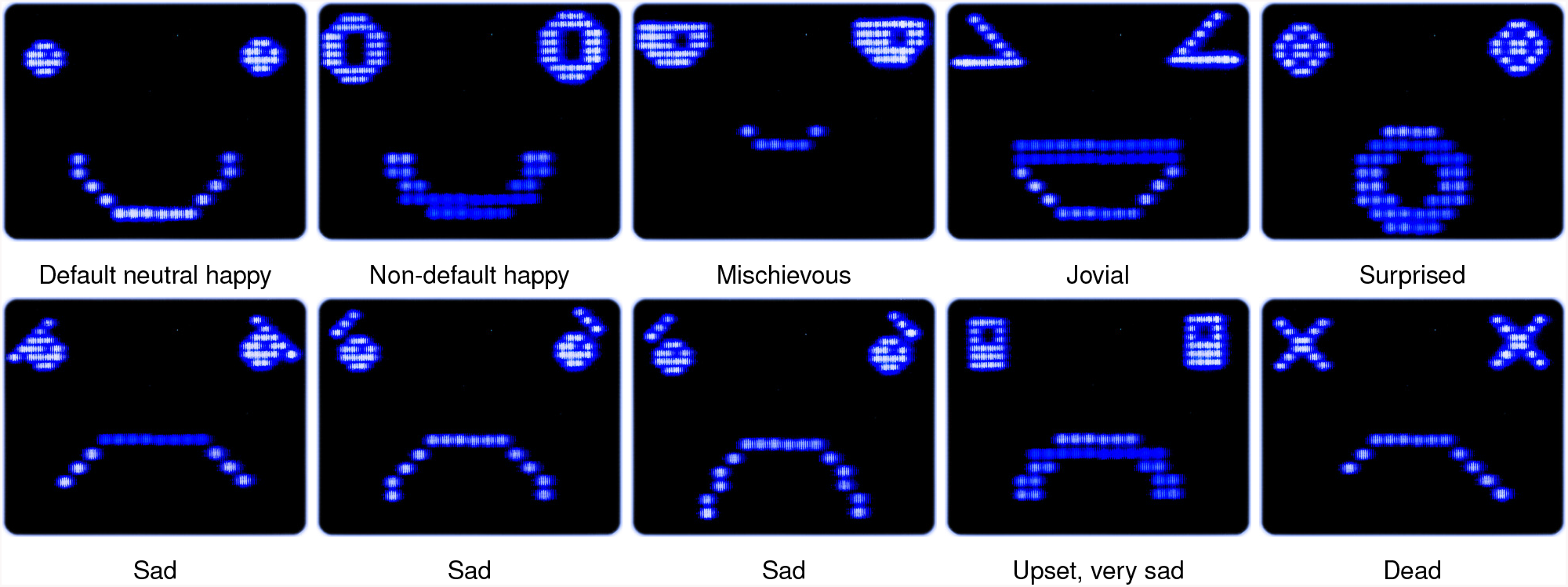
Faces which we found had clear expressions of emotion to subjects in our pilot study ^57^.

#### Demonstrations of Robot

Anecdotally, we have used the robot for demonstrations to multiple hundreds of children, typically functioning, as part of outreach efforts, and they often respond to the face. When the face changes from being happy to sad, there is often an audible “awww” from the audience, indicating that there is some empathetic connection with the robot or at the least an understanding of the emotion the robot is conveying.

### Overall Hardware and Software

We achieved a low cost system. The total cost of the system is much lower than our previous iteration. The motors cost about $400 USD, the computer cost a total of $600 USD, the battery cost $130 USD, the Kobuki base cost $500 USD, the pair of RealSense cameras cost $300 USD, the fisheye camera cost $45 USD, and the screen and controller cost $78 USD. There is also a cost for the various cables, primer, raw materials, and paint used. Many of the parts were either laser cut or 3D printed, which is not cheap. If these designs were to be fully translated, these components would be cheaper and higher volume production techniques would be used.

The hardware design and software integration effort which we have presented here represented a significant improvement over our prior design in many respects. We achieved a more robust system. The humanoid, which is the central component of the system is much more modular than our prior system and easier to maintain. In over a hundred hours of testing, its motors have never failed from over heating. However, the motors used suffer from communication challenges and in a future iteration should be upgraded. We would like to improve the motor and allow for more robust, faster, and multitasking communication over the USB. Another achievement is a long run time for experiments. The system easily runs for one and a half hours when all systems in the stack are running and an operator is connected. However, the current size of our internal storage limits how long the robot can capture data.

We have achieved a lightweight and portable system. Given the building materials, the system is light weight at only 9.8 kilograms and portable. It can be easily carried by one person, although its height can make it awkward. One future desire is to investigate the impact of the height on patient’s trust: would a shorter system may make the patient feel more confident or would being taller may make the patient more likely to listen to instructions ^58^.

The software stack we have developed enables telepresence and remote communication. The web interface does a good job of controlling the robot and demonstrating how easily such a system can be controlled. Although useful for rapid development, using ROSBridge to communicate over a network outside the robot is non-ideal, preventing scaling, redundancy, and limiting options for privacy. The system which we have could be easily ported by a web developer to use ROSBridge and NodeJS or similar within the robot to communicate securely with a remote server over the Internet.

Although the use of ROS presents some challenges for security and efficiency, it allows the system to be built upon and iterated. In our current implementation, there are limitations placed by available CPU time and storage space on new features. With further optimizations and ever improving computers, these challenges are addressable.

Driving can be challenging for a first time user, but the final set of camera positions does not make it overly onerous. For long term deployment, autonomous navigation would be preferred, the combination of ROS and the RGB+D cameras should make the addition of that functionality possible.

## Future Work

We are beginning to test the system with CP, BP, and typical subjects. Subjects play a Simon Says and target touch game in three conditions: the social robot with telepresence, the telepresence system without the social robot, and in person face to face with the operator. Subjects are assigned an order of conditions in a block random fashion. In all conditions the same games and script are used. Subjects are measured using a box and block test, grip strength test, and color trails test. Surveys and video grading, both automatic and manual, are being used to measure subject engagement, enjoyment, understanding, and compliance. The data which is collected will be used to help develop automated assessment tools.

There are a number of changes which we have planned for the system. We want to improve the mechanical design inside the head of the robot to make it more stable, improve the tint of the screen, and improve the finish of the head’s printed parts. We would like to eventually switch to using dynamixel motors within the arms to improve their controllability. Doing so would also make it easier to add lighting in the body and arms, which could run off the dynamixel bus. We would like to use a larger screen, which would also require a more capable base, and would likely come with an increase in system size and cost.

## Ethical Considerations

Increasing autonomy of robots in healthcare settings often raises ethical issues. In general there are questions about the biases implicit in data based algorithms that will eventually drive most robots ^59^. And some are concerned that robots will displace healthcare workers or simply be a distraction to care ^60^. However, failing to use robotic technologies to improve patient care could be considered irresponsible and in itself presents ethical concerns. The Lil’Flo robot described here melds telepresence, robotics, and computer vision to allow more patients to access healthcare while promoting patient-clinician interactions. In doing so, we hope to address some of the shortage of clinicians and caretakers in the rehab and care spaces. In view of the growing shortage of rehabilitation healthcare workers, we believe these technologies offer a solution to clinicians to help clinicians avoid compromising care as populations in need of care grow.

Beyond the ethics of care, in the current climate of growing concern over personal security and privacy there is concern that collecting, aggregating, and learning from large amounts of subject data could make some subjects uncomfortable and their data vulnerable to hackers. It is therefore imperative to both communicate clearly to subjects/patients what is being collected and how it is being used as well as taking every precaution to safeguard any data which we have. It is not however clear how to safely share data within the research community to accelerate development. In general, Human Subject Ethic Committees do not allow the publication of identifying data. To address this challenge, we ask all of our subjects for media and information releases above and beyond what is included in the standard consent. Subjects are welcome to participate in the study without agreeing to such releases. If they choose to opt-in, then their data will be compiled into a data set which can be released to other researchers to help accelerate development of novel algorithms. Balancing the competing needs of privacy and compliance against research progress can be difficult, but is important.

Discussions around the ethics in these spaces are growing as media coverage grows. It is important that roboticists and clinicians think about the ethical issues implied in the technologies which they are developing and using, try to understand the feelings of their subjects and patients, and be present in the public discussions on these issues.

## Conclusion

We have created a telepresence robot which can interact with patients in the communities where they live, learn, work, and play. To help bridge the gap between telepresence interactions and in person ones, we have attached a humanoid robot to our system. The humanoid can play games with patients and demonstrate movements to them, as a peer. This is designed to engage subjects and help them better understand the activities which they need to do, while keeping the clinician closely involved.

Compared to other systems which are available and described in the literature, we believe that our system is unique. It addresses the challenges which we set out to tackle and will enable us to examine how telepresence and social robotics can be used together for telerehabilitation.

Interested researchers can view the code to run the entire system at https://www.med.upenn.edu/rehabilitation-robotics-lab/lilflo-code. html and the mechanical design at https://www.med. upenn.edu/rehabilitation-robotics-lab/lilflo-cad.html.

## Data Availability

The code for the system is available on GitHub and the CAD is available on OnShape.

https://www.med.upenn.edu/rehabilitation-robotics-lab/lilflo-code.html

https://www.med.upenn.edu/rehabilitation-robotics-lab/lilflo-cad.html

## Declarations

### Guarantor

MJJ

### Contributorship

MJJ conceived the original idea for a telepresence-based social robot. MJS led the design, fabrication, and software integration of the system. VGL aided with the design and fabrication of the base. MJS led the composition of this manuscript with support from VGL and MJJ.

## Acknowledgements

Many people have contributed to this project. We want to especially thank Enri Kina and Carla Diana for their work on the design of the robot. We also want to thank the many clinicians who have contributed their opinions to the system, especially Dr. Laura Prosser and Dr. John Chuo.

